# A reference measurement of circulating ATPase inhibitory factor 1 (IF1) in humans by LC-MS/MS: comparison with conventional ELISA

**DOI:** 10.1101/2020.05.11.20097816

**Authors:** Annelise Genoux, Thibaut Duparc, Jean-Bernard Ruidavets, Cécile Ingueneau, Souad Najib, Jean Ferrières, Bertrand Perret, Mikaël Croyal, Laurent O. Martinez

## Abstract

ATPase inhibitory factor 1 (IF1) is a 9.5 kDa protein that binds to mitochondrial and plasma membrane ATP synthase and selectively inhibits ATP hydrolysis. Recently, IF1 was identified in systemic circulation in humans. IF1 appeared as an independent determinant of HDL-cholesterol with lower levels in coronary heart disease (CHD) patients. Moreover, IF1 was also found to negatively associate with mortality in these patients, supporting the notion that circulating IF1 could be a promising biomarker of cardiovascular disease. However, in previous studies, IF1 was quantified by a non-standardized competitive enzyme-linked immunosorbent assay (ELISA). Herein, we have validated a liquid chromatography–tandem mass spectrometry method (LC-MS/MS) enabling the accurate quantification of IF1 in human plasma. Plasma IF1 was trypsin-digested through an optimized procedure before LC-MS/MS analysis. The method was successfully validated over 4 independent experiments into the range of 100-1,500 ng/mL. Intra- and inter-assay variation coefficients had never exceeded 14.2% and accuracy ranged between 95% and 102% for the selected EAGGAFGK peptide marker. Subsequently, the results of the LC-MS/MS method were compared with those obtained using ELISA in 204 individuals from the GENES study. We found that IF1 plasma levels obtained using both techniques were strongly correlated (r =0.89, p <0.0001), while the Bland-Altman plot did not indicate any major statistically significant differences. To clinically validate LC-MS/MS, we confirmed the positive correlation between IF1 plasma levels and HDL-cholesterol (r =0.38, p <0.0001). Besides, we found lower IF1 plasma levels in CHD patients compared to controls (431±132 ng/mL and 555±173 ng/mL, respectively; p <0.0001). Hence, it can be concluded that the presented LC-MS/MS method provides a highly specific strategy for IF1 quantification in human plasma and could be proposed as a reference technique.

**Graphical abstract:** 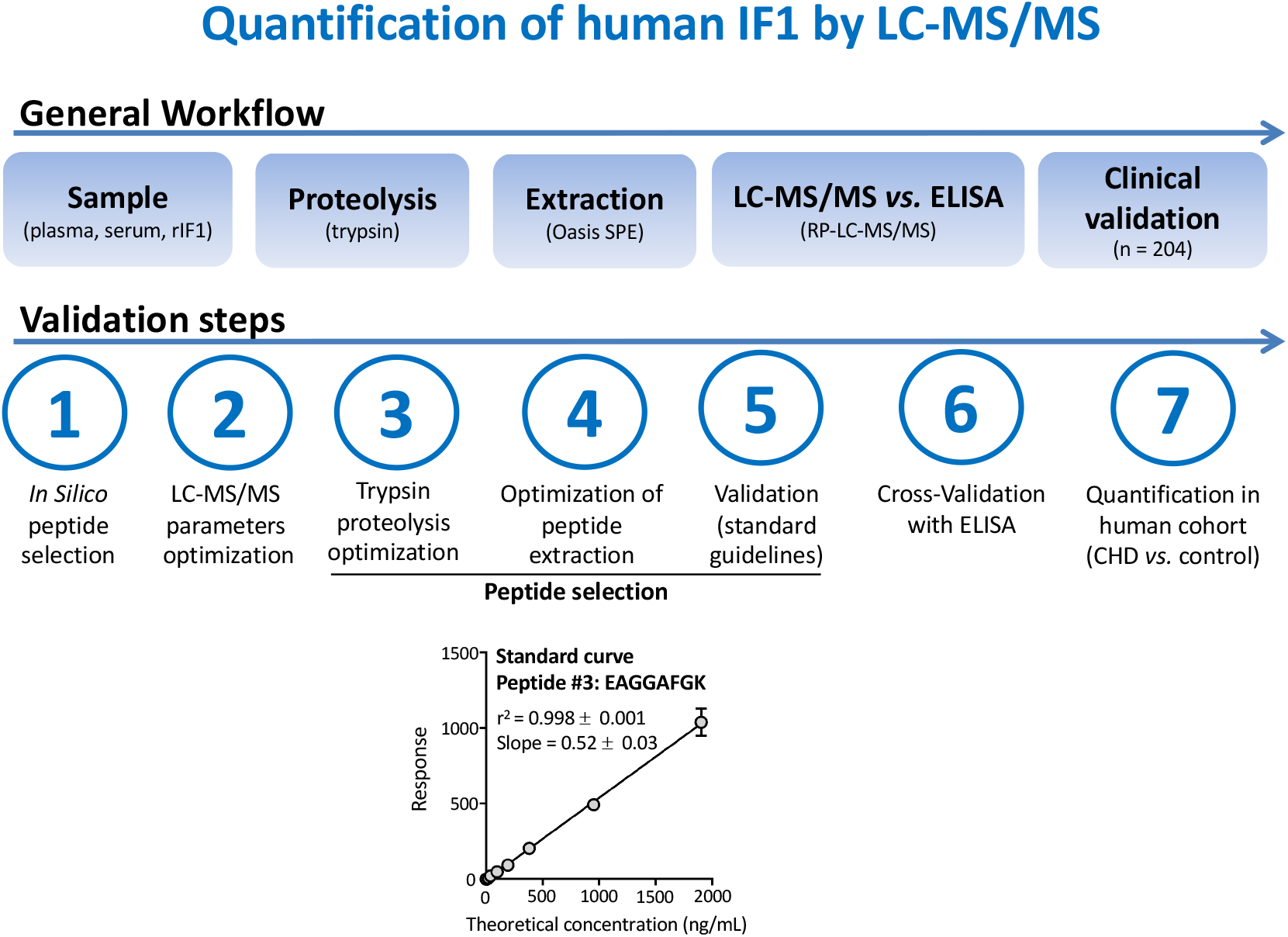

**Highlights:**

- ATPase inhibitory factor 1 (IF1) is a biomarker of coronary heart disease in humans.
- Thus far, IF1 plasma concentrations have been determined by non-standardized competitive ELISA.
- Here, we have developed and validated an LC-MS/MS assay for the accurate IF1 quantification in human plasma.
- Our LC-MS/MS assay is rapid and incurs reasonable costs, thus making it amenable for clinical practice.

## 1. Introduction

Despite many advances in therapeutics, cardiovascular diseases (CVD) remain the leading cause of deaths worldwide. Moreover, the main objective of current management guidelines for CVD is improving the prognosis in high-risk populations. Therefore, there is a critical need to determine novel non-invasive biomarkers to better predict the risk of CVD in the most at-risk patients.

Inhibitory Factor 1 (IF1) is a nuclear-encoded protein known to inhibit the ATP hydrolysis activity of both mitochondrial and plasma membrane ATP synthase. Consequently, IF1 prevents cell death in ischemic conditions and blocks endocytosis of high-density lipoprotein (HDL) by the liver [1,2]. In recent studies, IF1 was unexpectedly detected in the systemic circulation in humans. Moreover, in clinical studies, circulating IF1 was found to be an independent determinant of HDL-Cholesterol (HDL-C) levels and its mean concentration was lower in coronary heart disease (CHD) patients than in paired control individuals (430 ng/mL and 530 ng/mL, respectively; p <0.001). Later, IF1 levels were found to independently and negatively associate with mortality in CHD patients [3–5]. Recently, higher levels of circulating IF1 were also reported to be significantly associated with greater muscle mass [6]. These studies support the clinical potential of IF1 measurement for the assessment of cardiovascular risk in high-risk populations.

Further, the aforementioned studies have been performed using competitive enzyme immunoassays with different polyclonal anti-IF1 antibodies and recombinant IF1 protein [3,6]. However, such immunoassays are limited by batch size and batch-to-batch variability regarding both polyclonal antibodies and recombinant proteins batches. Thus, the use of a technique that will not depend on reagents’ availability and variability and will enable to establish a reference measurement of circulating IF1 for clinical practice is warranted. In this respect, enzymatic proteolysis of proteins by endoproteases, post-proteolysis concentration of peptides by solid-phase extraction (SPE) and subsequent targeted analysis of signature peptides by liquid chromatography-tandem mass spectrometry (LC-MS/MS), has been recently demonstrated as a powerful method to quantify low-abundance plasma proteins [7]. The aim of the present study was to develop and validate such an LC-MS/MS assay, enabling the quantification of circulating IF1 and its comparison to the traditional enzyme-linked immunosorbent assay (ELISA) approach.

## 2. Materials and Methods

### 2.1. Chemical and reagents

UPLC/MS-grade acetonitrile (AcN), water, methanol (MeOH), trifluoroacetic acid (TFA) and formic acid (FA) were purchased from Biosolve (Valkenswaard, Netherlands). Samples were prepared with the ProteinWorks™ eXpress kit (Waters Corporation, Milford, MA, USA), according to the manufacturer’s instructions and consisting in ammonium bicarbonate (50 mM, pH 8) as the digestion buffer, RapidGest (7 mg/mL in digestion buffer) as the detergent, dithiothreitol (70 mM in digestion buffer) as the reducing agent, iodoacetamide (142 mM in digestion buffer) as the alkylation agent, trypsin (7 mg/mL in HCl 1 mM) as the endoprotease, and 20% TFA as the inactivating agent. Moreover, the protein sequence of IF1 was BLAST searched using the UNIPROT tool (www.uniprot.org), and theoretical peptides were searched using ExPASy (http://web.expasy.org/peptide_mass) [8]. Synthetic labelled and unlabelled proteotypic peptides (**Table 1**) were provided by Thermo Scientific Biopolymers (Darmstadt, Germany). Stock solutions (1 mM) were prepared in 50% AcN containing 0.1% FA and stored at −20 °C until use. The mature human recombinant IF1 protein was chemically synthesized by GenScript (Piscataway, NJ, USA) at >80% purity. The human IF1 stock solution (1 mg/mL) was prepared in ultra-pure water and stored at −20 °C until use.

**Table 1.**
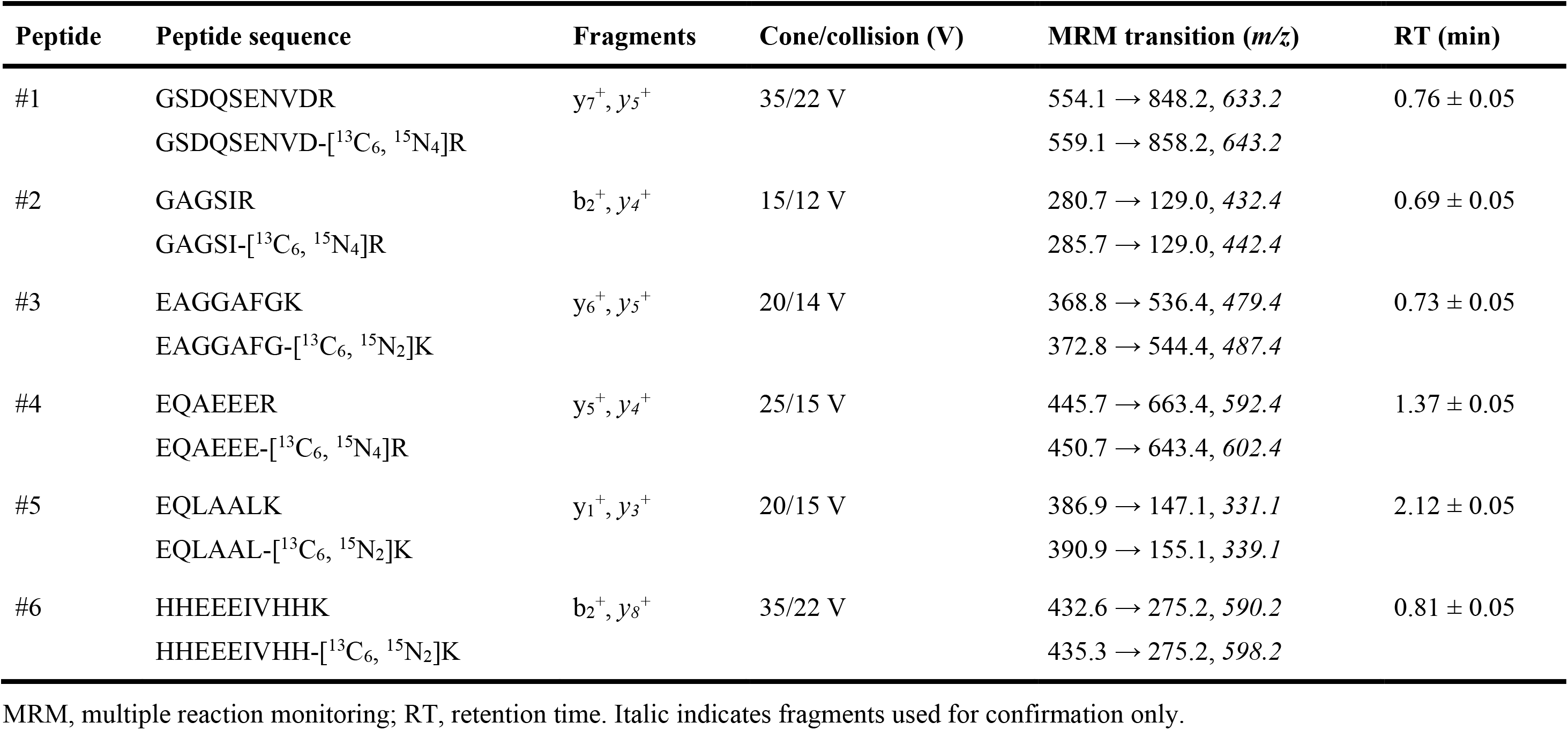
Summary of the analytical parameters selected for peptide detection.

### 2.2. Study samples

Ethylenediaminetetraacetic acid (EDTA) plasma and serum samples were obtained from 204 male participants from the case-control GENES (Génétique et Environnement en Europe du Sud) study [4]. The study protocol was approved by the local ethics committee (CCPPRB, Toulouse/Sud-Ouest, file #199-48, Feb 2000), with all patients having provided written informed consent. The sample collection has been declared as DC-2008-463 #1 to the Ministry of Research and to the Regional Health authority [4]. As previously described [4], the study’s participants (n = 99) were stable CHD patients, ages 45–74, living in the Toulouse area (southwest France) and prospectively recruited from 2001 to 2004 after admission to the Cardiology Department, Toulouse University Hospital, for cardiovascular examination and referred for evaluation and management of their CHD. During the same period, individuals of the same age range as the study participants but without CHD (further referred to as controls, n = 105) were selected from the general population using electoral rolls. Pools of human EDTA plasma and serum (8 donors; 50% males) were also purchased from the French Blood Bank (Nantes, France).

### 2.3. Standard samples and quality controls

Quality control (QC) samples were prepared at 4 concentration levels (0, 100, 600, 1500 ng/mL; i.e. 0, 10, 63, 158 nM). Concentrated solutions of human IF1 protein were prepared in water and then diluted 20-fold in pools of plasma (QC samples) or in water (control samples). In parallel, a mixed solution of unlabelled proteotypic peptides was constituted and serially diluted in water to obtain seven standard solutions, ranging from 200 to 10 nM (i.e. equivalent to 1,903-95 ng/mL of human IF1). A mixed solution of labelled peptides (10 μM) was prepared and added to the digestion buffer to create a final concentration of 100 nM.

### 2.4. Sample preparation

Samples were prepared with the ProteinWorks™ eXpress kit using an optimized procedure. Samples (40 μL) were incubated for 10 min at 80 °C in digestion buffer containing labelled peptides (100 μL) and RapidGest detergent solution (7 mg/mL, 10 μL), reduced for 20 min at 60 °C with dithiothreitol (20 μL), alkylated for 30 min at room temperature in the dark with iodoacetamide (30 μL), and digested for 8 hours at 37 °C with trypsin (30 μL). Enzymatic digestion was stopped with 20% TFA (5 μL). After 15 min at 45°C, precipitates were removed by centrifugation (15 min, 10 °C, 10,000 × g), and supernatants were cleaned using 30-mg Oasis HLB Cartridges (Waters Corporation), which were conditioned (100% MeOH; 1 mL), equilibrated (100% water; 1 mL), loaded (sample; ~200 μL), washed (5% MeOH, 0.1% TFA; 1 mL), and eluted (60% MeOH, 0.1% TFA; 500 μL). Eluates were dried under a gentle stream of nitrogen (45 °C), reconstituted with 10% AcN containing 0.1% FA (70 μL), and injected into the LC-MS/MS system.

### 2.5. Analytical parameters

Analyses were performed on a Xevo^®^ TQD mass spectrometer with an electrospray ionization (ESI) interface and an Acquity H-Class^®^ UPLC^TM^ device (Waters Corporation). Labelled and unlabelled peptides were separated on an Acquity^®^ BEH C_18_ column (1.7 μm. 2.1 × 100 mm, Waters Corporation) held at 60 °C with a linear gradient of mobile phase B (AcN containing 0.1% FA) in mobile phase A (5% AcN in water containing 0.1% FA) and at a flow rate of 500 μL/min. Mobile phase B was linearly increased from 1% to 70% for 5 min, kept constant for 1 min, returned to the initial condition over 1 min, and kept constant again for 1 min before the next injection. Subsequently, reconstituted sample (10 μL) were injected into the LC column. Labelled and unlabelled peptides were then detected by the mass spectrometer with the ESI interface operating in the positive ion mode [capillary voltage, 3 kV; desolvation gas (N_2_) flow and temperature, 1000 L/h and 450 °C; source temperature, 120 °C]. The multiple reaction monitoring (MRM) mode was applied for MS/MS detection. Selected MRM transitions, cone voltages, and collision energies are described in **Table 1**.

### 2.6. Optimization of sample preparation

The method was developed and optimized using QC samples spiked with known amounts of human IF1 protein. For trypsin proteolysis, 8 incubation times (0, 1, 2, 3, 4, 8, 16 and 24 hours) were assessed (*n* = 6). Solid-phase extraction (SPE) was also optimized from QC samples treated as described above. The wash and elution conditions were optimized with increasing levels of MeOH or AcN in water, with or without additives (0.1% TFA or 0.5% NH_4_OH), added directly after sample loading (*n* = 3). Several mixtures of AcN in water containing 0.1% FA were also tested to reconstitute sample after drying of peptide solutions at 50 nM (*n* = 3).

### 2.7. Method validation

The LC-MS/MS assay was validated *via* four independent experiments with two calibration curves and 24 plasma QCs (4 concentration levels; *n* = 6). The assay linearity was illustrated by R^2^ coefficients calculated from calibration curves by linear regression analysis (1/× weighting, origin excluded). The intra- and inter-assay variability was expressed by coefficients of variation (CVs) obtained at each QC level. The assay accuracy was expressed by the mean bias between the theoretical and measured concentrations at each QC level after baseline subtraction (i.e., the mean measured concentration of unspiked QC). The targeted lower limit of quantification (i.e. 100 ng/mL of IF1) was validated with a signal-to-noise ratio greater than 10 and a variability and accuracy lower than 20%. Limits of acceptance were set at ± 15% for accuracy and variability at other concentration levels [9]. Cross-validation was performed by comparing results obtained by standard ELISA and LC-MS/MS on plasma samples from 204 subjects. For quantitative experiments, plasma samples spiked with 5 nM (375 ng/ml) of human IF1 (*n* = 3) or digestion buffer (*n* = 3) were used to check the completion of the enzymatic reaction and the reliability of the assay.

### 2.8. Data management and statistical analysis

IF1 protein concentrations were calculated using calibration curves plotted from standard solutions and expressed in nM, assuming that 1 mole of peptide was equivalent to 1 mole of protein. The concentrations were then converted to standard units (ng/mL) using the IF1 molecular weight (9,517 g/mol). Data are presented as percentages for qualitative variables and as means with standard deviations (SD) for quantitative ones. The Shapiro-Wilks test was used to test the normality of distribution of residuals. A Spearman correlation test and a Bland-Altman plot were generated to compare both methods [10]. For the Bland-Altman plot, the difference of values (y-axis) obtained from the two methods was plotted according to the average value obtained with the two methods. The mean difference and the limits of agreement, corresponding to the 95% confidence level (i.e., mean ± 1.96 × SD), were calculated. The degree of correlation and agreement between measurements was assessed by calculating the intra-class correlation coefficient (ICC) [11]. Comparison between groups was made using two-tailed unpaired Student *t*-test. Spearman coefficients were calculated to study the correlations between IF1 and HDL-C. Outcomes of p <0.05 were considered as statistically significant. Graphics and statistical analyses were achieved with GraphPad Prism software (version 6.0, GraphPad Software Inc., La Jolla, CA, USA).

## 3. Results and Discussion

### 3.1. Selection of peptide markers

*In silico* investigations led to the identification of 6 candidate peptides able to specifically target the human mature IF1 protein. Peptides and their labelled internal standards were thus synthesized for optimization of the MRM transitions used for LC-MS/MS analyses (**Table 1**). Candidate peptides were primarily detected as doubly charged precursor ions. After MS/MS fragmentation, each precursor ion yielded several specific and singly charged “y” (C-term) or “b” (N-term) product ions, thereby ascertaining the peptide sequences. For each candidate peptide, the most intense product ion was selected to quantify human IF1 protein levels. Moreover, the product ion second in intensity was kept to confirm the specific detection of the target peptide in biological samples.

### 3.2. Optimization of sample preparation

Optimization of trypsin proteolysis showed contrasting released profiles between candidate peptides (**Fig. 1A**). Contrary to manufacturer’s recommendations (2h, 45 °C), the optimal condition was reached with a trypsin incubation at 37 °C for 8 h. However, only two peptides were satisfactorily generated (#3: EAGGAFGK; #5: EQLAALK). Our further investigations revealed that peptide #1, peptide #2, peptide #4 and peptide #6 displayed several deleterious properties, including a more pronounced matrix effect, a poor stability during sample preparation, and a poor ionization yield, as compared to peptide #3 and peptide #5. Hence, these peptides were not considered for the assay validation. All other manufacturer’s recommendations (linearization, reduction, and alkylation) were confirmed by testing four incubation times on the pooled human plasma (not shown). Moreover, SPE was also optimized on plasma samples spiked with known amounts of recombinant IF1 (QC). Moreover, MeOH was slightly more efficient than AcN for peptide recovery, and an acidic additive (TFA) markedly improved extraction by up to 40%. The optimized wash and elution solvents used were 5% and 60% methanol containing 0.1% TFA, respectively (**Fig. 1B**). After drying, several mixtures were tested for sample reconstitution to obtain the optimal signal intensity. The optimal mixture was determined to be 10% acetonitrile containing 0.1% TFA for peptide #3 and peptide #5 (**Fig. 1C**). Higher concentrations of acetonitrile led to peak distortions and signal loss. Despite SPE, plasma strongly reduced peptide detection by ~25%, compared with aqueous controls. These matrix effects were corrected after internal standard normalization. Of note, serum was also tested and showed higher signal loss (~60%) with a more pronounced variability than that observed with plasma. Different plasma volumes (10, 20, 40, and 50 μL) were also assessed and 40 μL was found to be the most suitable for assay sensitivity and proteolysis efficiency (not shown).

**Figure 1.**
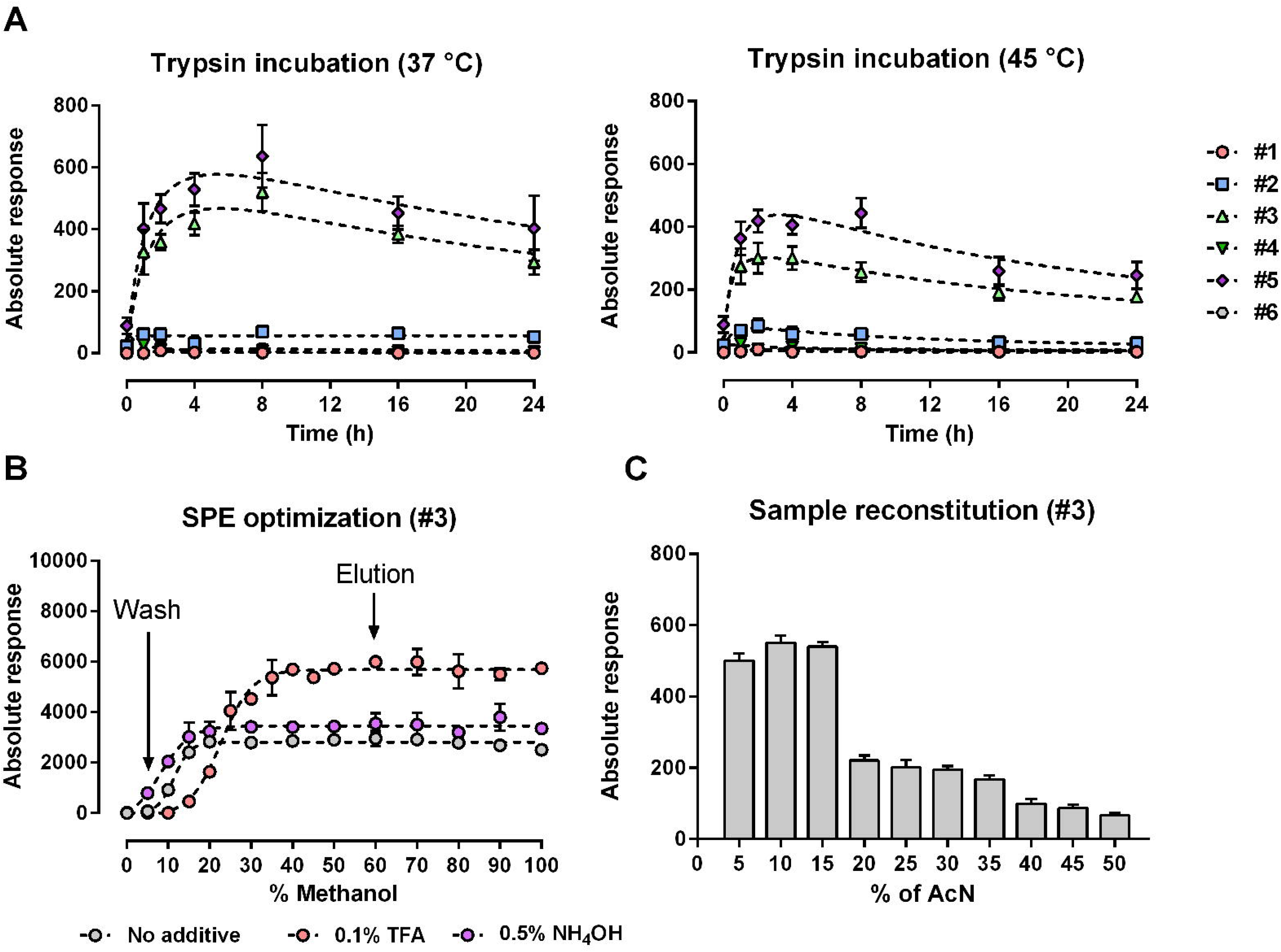
Optimization of sample preparation for IF1 quantification. **(A)** Trypsin incubation times were optimized using a mixture of human plasma spiked with a known amount of human IF1 (600 ng/mL, *n* = 6). **(B)** Solid-phase extraction protocols were optimized using a mixture of human plasma spiked with a known amount of human IF1 (1,500 ng/mL, *n* = 3, peptide #3). **(C)** Solvent composition for sample reconstitution of dried samples was optimized by assessing various concentrations of acetonitrile (AcN) in water with 0.1% FA (*n* = 3, peptide #3). The values represent means ± standard deviations.

### 3.3. Method validation

To test the linearity of the method, a regression model (1/× weighting and origin excluded) was used. After analysis of the 8 calibration curves (2 calibration curves per experiment), the mean *R^2^* value were 0.998 ± 0.002 and 0.996 ± 0.001 for peptide #3 and peptide #5, respectively, and the CVs never exceeded 9.1% over the concentration range tested (**Table 2**). Of note, at the targeted lower limit of quantification (95 ng/mL of human IF1), the signal-to-noise ratios were of 23 ± 4 and 7 ± 2 for peptide #3 and peptide #5, respectively. Human IF1 was also found to remain stable for 6 months at −80 °C and after 3 freeze/thaw cycles (−80 °C) in human EDTA plasma. Stock solutions of the synthetic peptides #3 and #5 were also stable for 6 months at −20 °C. Finally, digested samples were found stable for 5 days in the refrigerated autosampler (10 °C) and 6 months at −20 °C (not shown). Intra- and inter-assay variability parameters were then assessed using calibration curves and QC samples. For peptide #3, intra- and inter-assay CVs did not exceed 14.2% and the mean measured concentrations did not deviate by more than 4.9% (mean bias) as compared to expected concentrations (**Table 2**). In contrast, intra- and inter-assay CVs were out of our acceptance criteria for peptide #5 (± 15%) [8,9]. These results indicate that solely peptide #3 (EAGGAFGK) can be used to accurately quantify human IF1 in plasma samples, and was thus retained for further analyses.

**Table 2.** Validation of the LC-MS/MS assay. The analytical validation was performed over four distinct experiments. Assay linearity was illustrated by the R^2^ coefficients and slopes (mean ± SD, *n* = 8) calculated from calibration curves by linear regression analysis (origin excluded, 1/× weighting). Intra- and inter-assay variability was expressed by the coefficients of variation (CVs) obtained at each QC level (*n* = 6 × 4 per level). Assay accuracy was expressed by the mean bias between the theoretical and measured concentrations after baseline subtraction (*n* = 6 x 4 per level). Stabilities were evaluated on QC samples stored at −80 °C and assayed with fresh calibration curves (*n* = 6 × 4).

| Parameters | #3: EAGGAFGK | #5: EQLAALK |
| --- | --- | --- |
| **Linearity** |  |  |
| $R^2$ | $0.998 \pm 0.002$ | $0.996 \pm 0.001$ |
| *Slope* | $0.52 \pm 0.03$ | $0.44 \pm 0.04$ |
| **Baseline (302 ng/mL)** |  |  |
| $CV_{\text{Intra}}$ (%) | 5.2 | 12.3 |
| $CV_{\text{Inter}}$ (%) | 7.1 | 20.1 |
| *Accuracy* (%) | N/A | N/A |
| **Low level (100 ng/mL)** |  |  |
| $CV_{\text{Intra}}$ (%) | 13.2 | 20.2 |
| $CV_{\text{Inter}}$ (%) | 14.2 | 29.2 |
| *Accuracy* (%) | $99.2 \pm 3.4$ | $112.3 \pm 23.2$ |
| **Middle level (600 ng/mL)** |  |  |
| $CV_{\text{Intra}}$ (%) | 4.2 | 12.1 |
| $CV_{\text{Inter}}$ (%) | 8.3 | 15.2 |
| *Accuracy* (%) | $95.1 \pm 3.4$ | $103.2 \pm 12.1$ |
| **High level (1,500 ng/mL)** |  |  |
| $CV_{\text{Intra}}$ (%) | 3.1 | 6.2 |
| $CV_{\text{Inter}}$ (%) | 3.7 | 7.1 |
| *Accuracy* (%) | $102.4 \pm 1.5$ | $92.4 \pm 7.2$ |
| **Stability** |  |  |
| *6 months* ($-80^\circ\text{C}$) (%) | $92.3 \pm 4.5$ | $89.2 \pm 4.7$ |
| *3 freeze/thaw cycles* (%) | $99.2 \pm 2.1$ | $92.1 \pm 4.3$ |

N/A: not applicable.

### 3.4. Comparison of LC-MS/MS with ELISA

For method comparison, we used 204 human plasma samples from the GENES study, in which IF1 levels were previously determined by the ELISA assay [4]. As shown in **Fig. 2A**, IF1 concentrations measured by LC-MS/MS from peptide #3 (*n* = 204) were significantly correlated with those obtained by ELISA (r =0.89, p <0.001). Intraclass correlation coefficient was 0.93 [95% CI, 0.91-0.95], indicating an excellent degree of correlation and agreement between LC-MS/MS and ELISA measurement. In the Bland-Altman test, the mean of both concentrations obtained by LC-MS/MS and ELISA was calculated and plotted against the difference between both measurements. The mean difference and limits of similarity, corresponding to the 95% confidence interval, were drawn and it was demonstrated that 195 out of the 204 values were within these limits (**Fig. 2B**). However, the Bland-Altman plot highlighted non-negligible concentration differences between the analytical approaches, with deviations of up to 50%, but only 9 outliers were identified (< 5%). After exclusion of these outliers, the mean deviation between both approaches was 3.4 ± 13.7 %. Hence, the human IF1 protein concentrations measured by LC-MS/MS were considered similar to those obtained by ELISA.

**Figure 2.**
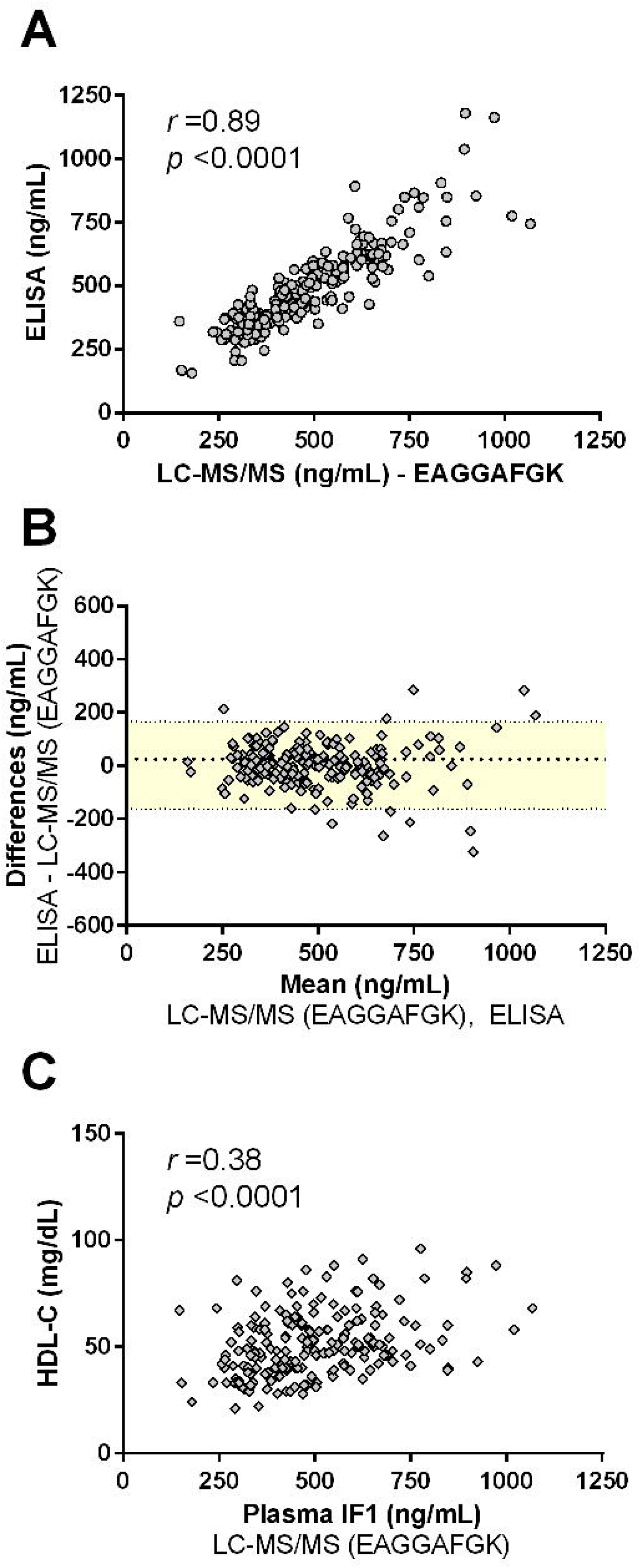
Comparison of the LC-MS/MS assay with ELISA in 204 individuals. **(A)** Spearman correlations obtained between ELISA and LC-MS/MS for IF1 serum concentration measurements. **(B)** Comparison of both methods using the Bland-Altman test. Bland-Altman plots were generated to test the similarity of methods. For the Bland-Altman plots, the difference of values (y-axis) obtained from the two methods was plotted according to the average value obtained by them. The mean difference and the limits of agreement (colored area), corresponding to the 95% confidence level (i.e., mean ± 1.96 × SD), are represented. **(C)** Spearman correlations obtained between IF1 plasma concentrations measured by LC-MS/MS and HDL-Cholesterol (HDL-C) levels.

### 3.5. Clinical validation

One of the main features of circulating IF1 is that its levels, measured by ELISA assay, were positively correlated with levels of HDL-C (r = 0.40, p< 0.001) [4]. Accordingly, from the Spearman correlations analysis, plasma IF1 measured by LC-MS/MS with peptide #3 was found to positively correlate with HDL-C (r = 0.38, p< 0.0001, **Fig. 2C**). Another important clinical output of measuring circulating IF1 level is that it was found to be significantly lower in CHD patients as compared to control individuals [4]. In the present cohort, the mean IF1 concentrations were found significantly lower in CHD patients than in controls when measured by LC-MS/MS (431 ± 132 ng/mL and 555 ± 173 ng/mL, p <0.0001) and ELISA (431 ± 118 ng/mL and 558 ± 180 ng/mL, p <0.0001). Altogether, these results confirmed that the LC-MS/MS method we have developed as part of the present study is clinically relevant for the accurate quantification of plasma IF1 levels in human samples.

### 3.6. Towards IF1 isoforms and polymorphisms

In humans, IF1 is a nuclear encoded protein translated as a precursor protein that undergoes the removal of the N-terminal 25-residue pre-sequence when imported to the mitochondria. At this stage, it is still unknown whether IF1 is circulating as the precursor or mature form. In this respect, our study does not allow to discriminate these forms since all selected peptide markers were present in the mature sequence of IF1.

Three IF1 isoforms, resulting from alternative splicing at *ATPIF1* gene locus, have been described in humans [3]. IF1 variant 1 (NCBI Reference Sequence: NP_057395.1) encodes the longest isoform, while IF1 variant 2 (NCBI Reference Sequence: NP_835497.1) and variant 3 (NCBI Reference Sequence: NP_835498.1) have a shorter and distinct C-terminus compared to isoform 1. The selected peptide #3 used in the present LC-MS/MS method is located within a sequence common to the three variants and thus enables the quantification of the 3 isoforms together. Among the candidate peptide markers (**Table 1**), peptides #5 and #6 are specific of isoform 1. However, peptide #6 was not efficiently released after trypsin proteolysis and validation parameters for peptide #5 were not within our acceptance criteria (**Table 2**). Hence, further studies in developing a methodology using one of these peptides will be warranted to specifically quantify isoform 1. However, a recent study performed in several human tissues reported negligible mRNA expression for isoforms 2 and 3 compared to isoform 1 [12], suggesting that isoform 1 is likely to be the major circulating isoform. Additionally, previous studies demonstrating the clinical interest of IF1 measurement in the circulation have been performed using polyclonal antibodies in competitive ELISA assays that did not differentiate between various IF1 isoforms [3,6], thus indicating that the clinical significance of IF1 is measurable with a method that does not specifically quantify isoform 1.

Regarding the current knowledge on the potential clinical impact of IF1 polymorphisms, we found that the single nucleotide polymorphism rs748968212 (Highest population Minor Allele Frequency (MAF) <0.01%, Ser39 > Phe39) could affect the phosphorylation state of IF1 and modulate its inhibitory activity on ATP synthase [13]. Interestingly Ser39 is located in the candidate peptide marker #2 (**Table 1**). In the future, developing a methodology using this peptide might serve to quantify circulating levels of IF1 in heterozygous patients for this mutation, as it has been previously developed for other isoforms of circulating proteins, such as apoE, for which production or catabolic rates are differently affected according to the specific isoforms [14].

## 4. Conclusion

Here, we have developed and validated a simple and fast LC-MS/MS assay enabling the accurate quantification of human IF1 in plasma samples into the range 100-1,500 ng/mL (i.e. 10-160 nM). Unlike most conventional LC-MS/MS based assays for low-abundancy proteins, protein concentration by immunoaffinity or immunodepletion was not necessary for our assay. Of note, LC-MS/MS allows a high level of multiplexing, enabling simultaneous quantification of several proteins and providing further information on a patient’s metabolic profile. In that respect, we have already described a validated LC-MS/MS assay for the quantification of several apolipoproteins in a single run [8,15]. Here, the proposed method was multiplexed for the simultaneous analysis of IF1 and major HDL-related apolipoproteins such as apoA-I, apoA-II, apoE, apoD and apoM ([8] and data not shown), thus making it amenable for a wide range of studies aiming to decipher the role of IF1 in CHD. Finally, the LC-MS/MS method developed in our study could be used in the future to determine IF1 turnover *in vivo* by measuring the incorporation of an injected labelled tracer, such as ^2^H_3_-leucine, over time, thus allowing the determination of its kinetic parameters, including production and catabolic rates [14].

## Data Availability

All data referred to in the manuscript are available upon request

## Authorship contribution statement

Conception and design of the study: A.G., M.C., L.O.M.; Methodology and validation: A.G., J.B.R., M.C.; Acquisition of data, or analysis and interpretation of data: A.G., T.D., J.B.R., C.I., S.N., M.C., L.O.M.; Drafting the article or critical revision of its content: A.G., J.F., B.P., M.C., L.O.M.; Funding acquisition: L.O.M. All authors have read and approved the final version of the manuscript.

## Acknowledgements

We are grateful to the Biogenouest Corsaire core facility for their financial support.

**Abbreviations**
AcN: acetonitrile
CHD: coronary heart disease
CI: confidence interval
CV: coefficient of variation
CVD: cardiovascular diseases
EDTA: thylenediaminetetraacetic acid; electrospray ionization, ESI
ELISA: enzyme-linked immunosorbent assay
FA: formic acid
HDL: high-density lipoprotein
IF1: inhibitory factor 1
LC-MS/MS: liquid chromatography–tandem mass spectrometry method
MeOH: methanol; multiple reaction monitoring, MRM
QC: quality control
SPE: solid-phase extraction
TFA: trifluoroacetic acid

